# Development and prognostic validation of a three-level NHG-like deep learning-based model for histological grading of breast cancer

**DOI:** 10.1101/2023.02.15.23285956

**Authors:** Abhinav Sharma, Philippe Weitz, Yinxi Wang, Bojing Liu, Johan Hartman, Mattias Rantalainen

**Affiliations:** Department of Medical Epidemiology and Biostatistics, Karolinska Institutet, Stockholm, Sweden; Division of Precision Medicine, Department of Medicine, NYU Grossman School of Medicine, New York, NY; Department of Oncology-Pathology, Karolinska Institutet, Stockholm, Sweden; Department of Clinical Pathology and Cancer Diagnostics, Karolinska University Hospital, Stockholm, Sweden; MedTechLabs, BioClinicum, Karolinska University Hospital, Solna, Sweden

**Keywords:** breast cancer, pathology, deep learning, image analysis, clinical decision support

## Abstract

Histological Grade is a well-known prognostic factor that is routinely assessed in breast tumours. However, manual assessment of Nottingham Histological Grade (NHG) has high inter-assessor and inter-lab variability, causing uncertainty in grade assignments. To address this challenge, we developed and validated a three-level NHG-like deep learning-based histological grade model. The primary performance evaluation focuses on prognostic performance.

This observational study is based on two patient cohorts (SöS-BC-4, N=2421 (training and internal test); SCAN-B-Lund, N=1262 (test)) that include routine histological whole slide images together with patient outcomes. A Deep Convolutional Neural Network (CNN) model with an attention mechanism was optimised for the classification of the three-level histological grading (NHG) from hematoxylin and eosin-stained WSIs. The prognostic performance was evaluated by time-to-event analysis of Recurrence-free survival (RFS) and compared to clinical NHG grade assignments in the internal test set as well as in the fully independent external test cohort. We observed effect sizes (Hazard Ratio) for grade 3 vs 1, for the conventional NHG method (HR=2.60 (1.18-5.70 95%CI, p-value = 0.017)) and the deep learning model (HR = 2.27, 95%CI: 1.07-4.82, p-value = 0.033) on the internal test set after adjusting for established clinicopathological risk factors. In the external test set, the unadjusted HR for NHG 1 vs 2 was estimated to be 2.59 (p-value = 0.004) and NHG 1 vs 3 was estimated to be 3.58 (p-value < 0.001). For predGrade, the unadjusted HR for grade 1 vs 2 HR=2.52 (p-value = 0.030), and 4.07 (p-value = 0.001) for grade 1 vs 3. In multivariable analysis, HR estimates for neither NHG nor predGrade were found to be significant (p-value >0.05). We tested for differences in HR estimates between NHG and predGrade in the independent test set, and found no significant difference between the two classification models (p-value > 0.05), confirming similar prognostic performance between conventional NHG and predGrade.

Routine histopathology assessment of NHG has a high degree of inter-assessor variability, motivating the development of model-based decision support to improve reproducibility in histological grading. We found that the proposed model provides similar prognostic performance as NHG. The results indicate that deep CNN-based models can be applied for breast cancer histological grading.

## Introduction

Histological grading is a well-established prognostic factor for breast cancer and is associated with the aggressiveness of the tumour (1). An assessment of three morphological features determines the histological grade of breast tumours. These features include tubular formation (glandular differentiation), nuclear pleomorphism, and mitotic counts, and each component is given a score from I to III. The sum of the sub-component scores enables the assignment of tumours into three grades (Grade 1-3), referred to as Nottingham Histologic grade (NHG), where Grade 1 is associated with a good prognosis and Grade 3 is associated with a poor prognosis (2).

However, the assessment of histological grading has a high inter-observer variability including the assessments of individual subcomponents of histological grading (3–5). A recent nationwide study in Sweden reported significant inter-lab variabilities for histological grading across different pathology labs (6). Such variabilities indicate an intrinsic uncertainty in routine NHG assessment and potential errors, which can cause both under and over-treatment of breast cancer.

Recent advances in high-resolution digital whole slide images (WSIs) have greatly enhanced the computer-based pathology workflow, paving the way to novel digital decision support solutions. Recently, deep learning-based analyses on WSIs have shown promising results in a multitude of tasks, including cancer classification, grading, and predictions of genetic mutations in prostate and lung cancers (7–9).

Deep learning, especially deep convolutional Neural Networks (CNNs), has been proven to be effective for modelling of WSI data, including in the application of breast cancer histological grading. Previously models for the classification of grades 1 and 2 (together) vs. grade 3 have been implemented for breast cancer (10,11). Jaroensri *et al*. implemented a model that classified the sub-components, and the sub-component score, for breast cancer histological grading and the prognostic performance was compared against routine classification (12). Wang et al. developed a model based on histological grade morphology in breast cancer that was applied to improve risk stratification of intermediate-risk patients (histological grade 2) (13).

To our knowledge, this is the first study focusing on the development of a deep-learning-based breast cancer histological grade classification with a three-level grading system resembling the routine NHG in breast cancer with prognostic evaluation. We evaluate the proposed model from the perspective of prognostic performance (time-to-event) in both internal test data and a fully independent external test cohort and compare it with the routine clinical grade assignment.

## Methods

### Study materials

The patients in this study were from two Swedish cohorts, SöS BC-4 (n = 2421), and the SCAN-B cohort (n = 1262). SöS BC-4 is a retrospective observational study that included patients diagnosed at Södersjukhuset (South General Hospital) in Stockholm between 2012 and 2018 that had archived histological slides available and also available histological grade information. Patients that had received neoadjuvant therapy were excluded. The SCAN-B cohort, which we used as an independent external test set, includes a subset of patients (n = 1262) enrolled in the prospective SCAN-B study (14), diagnosed between 2010 and 2019 in Lund, Sweden. Both cohorts consist of female patients diagnosed with primary invasive breast cancer. Patients’ clinical information (i.e. clinical NHG, estrogen receptor status, Her2 status, tumour size, and lymph node status) was retrieved from the Swedish National Registry for Breast Cancer (NKBC). Whole Slide Images (WSIs) were generated (40X magnification) using Hamamatsu NanoZoomer histopathology slide scanners (S360 or XR) from clinical routine H&E-stained, formalin-fixed paraffin-embedded (FFPE) resected tumour slides. We included one H&E WSI per patient, which was either the established primary diagnostic fraction or otherwise the H&E WSI with the largest predicted tumour area. The CONSORT diagram for this study is provided in Figure 1.

**Figure 1.**
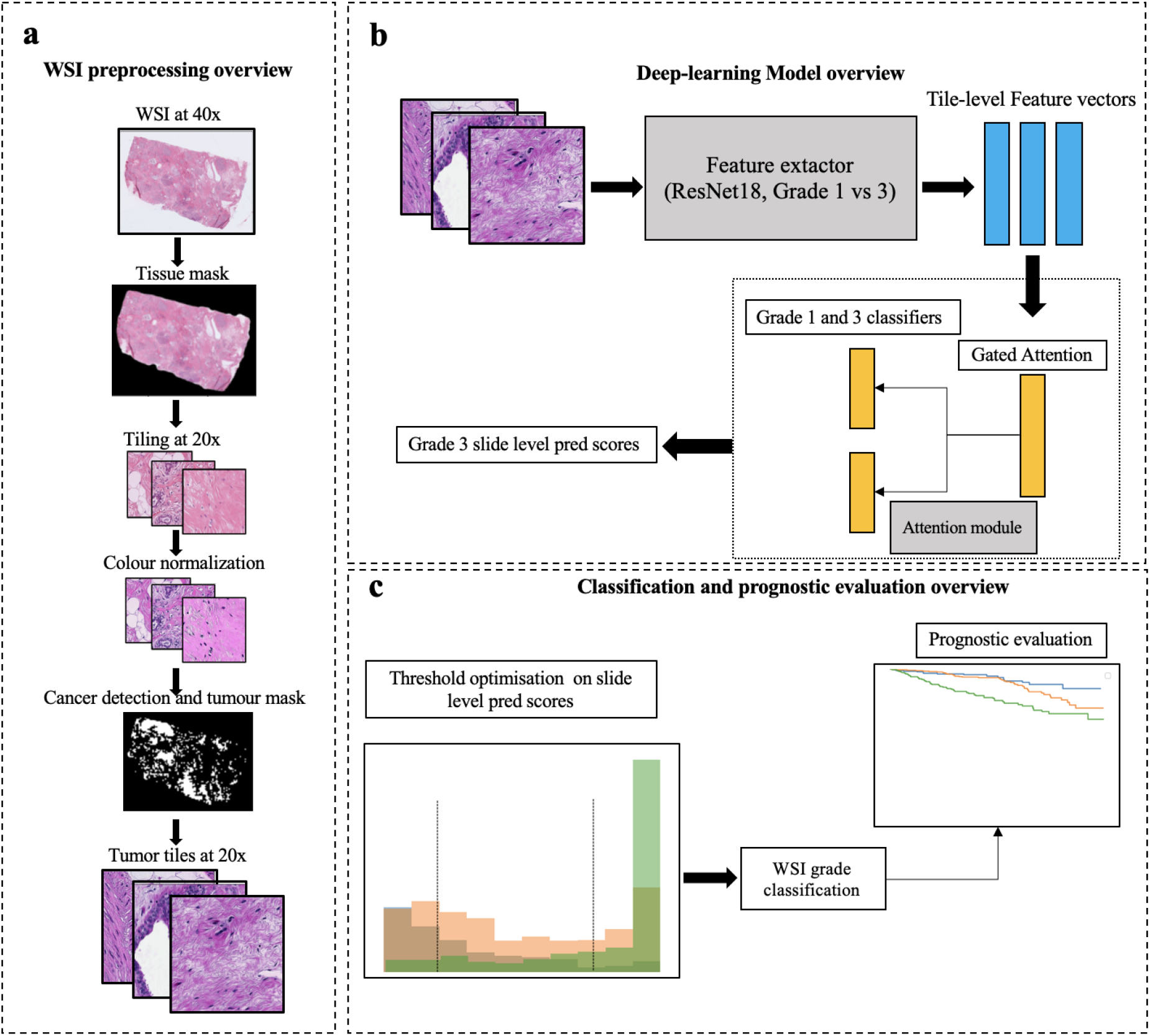
Overview of the image pre-processing, model optimisation and performance evaluation. **a**. Standardised WSI preprocessing pipeline from retrieval of WSI at 40x magnification to the cancer-detected tumour tiles from the WSI. **b**. Schematic overview of the image modelling strategy, including the deep CNN feature extractor and attention module. Model optimisation, hyperparameter tuning and model selection were performed by cross-validation (CV). In each CV training round, the Feature extractor and Attention module were trained from cancer tiles in the CV training set. In each CV validation round, the features extractor and attention model were re-optimised and subsequently, the CV validation set was evaluated. **c**. Two cut-offs were further derived from the slide-level prediction scores, which categorised the prediction scores into three-level predicted grades. The cut-offs were optimised by maximising the agreement between the predicted grade and clinical NHG. We further evaluated the prognostic performance of the predicted grade on recurrence-free survival.

### Image pre-processing and deep learning modelling methods

WSIs were pre-processed and quality controlled in a standardised processing pipeline, followed by model optimisation and performance validation of the system (Figure 1a).

### WSI Preprocessing

The WSI pre-processing pipeline has been previously described in detail in (13). A brief overview of the preprocessing steps is shown in Figure 2. First, we generated tissue masks excluding most of the backgrounds from the WSIs. We added a maximum value of 25 on the Otsu threshold in order to reduce the removal of the tissue regions in some cases due to the high threshold value on the transformed saturation channel. The tissue regions were divided into image patches (i.e. tiles) of size 1196 × 1196 pixels. The image tiles were down-sampled by a factor of two from the original scanning resolution (40X) to 20X resolution (598 × 598 pixels; 271 × 271 *µ*m). Next, we applied the Laplacian filter (OpenCV package version 3.4.2) on all the image tiles and computed the variance of the filtered tiles. Tiles with a variance lower than 500 units were considered blurry and excluded from further analyses (15) To mitigate stain colour variability, the colour normalization method described by Macenko et al. (16) was applied, with a modification to enable WSI-level colour correction, as previously described in (13). Lastly, we applied a pre-trained CNN model developed in (13) to detect invasive cancer in our current study population. Only tiles predicted as invasive cancer from the pre-trained model were considered as regions of interest and thereafter included in further analyses.

**Figure 2.**
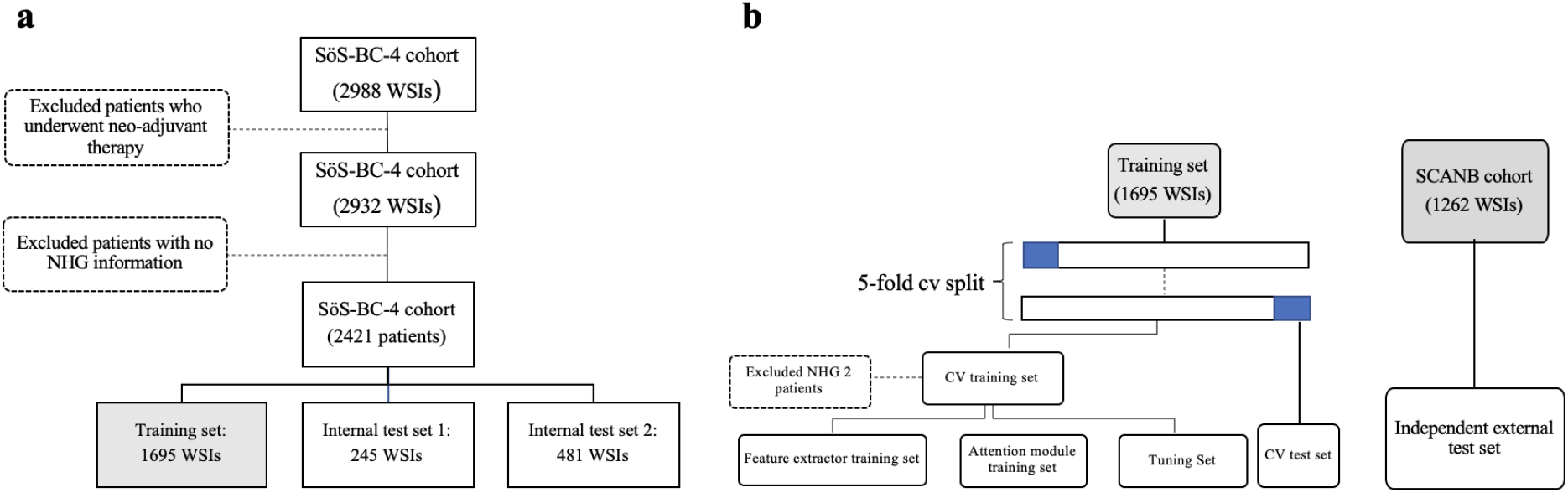
SöS cohort splitting criteria. **a**. The SöS cohort was first split into the training, internal test set 1, and internal test set 2 on the patient level. The split was stratified by clinical histological grading (NHG), estrogen receptor (ER) status, epidermal growth factor receptor 2 (HER2), and Ki-67 status. **b**. A five-fold cross-validation (CV) split was further generated on the patient level within the training set (n=1695 WSIs). Each CV fold consisted of a CV training set (80%) and a CV test set (20%) balanced on clinical NHG. The CV training set is further sub-split into the Feature extractor training set (50%), the Attention module (40%), and the Tuning set (10%)

### Image analysis using deep learning

The SöS cohort was used for model development and internal validation. The cohort was split into the training set (n = 1695), internal test set 1 (n = 245 WSIs), and internal test set 2 (n=481 WSIs) as shown in Figure 2a). The training and internal test sets were split on the patient level and stratified by histological grading (NHG), estrogen receptor (ER) status, epidermal growth factor receptor 2 (HER2) and Ki-67 status.

The training and optimisation of the feature extractor and attention module were performed on the training set (n=1695) using five-fold cross-validation (CV). For each CV-fold, the training set was split into a CV training set (80%) and a CV test set (20%) stratified by histological grading (NHG) as shown in Figure 1 b). The CV training set was further sub-split into the Feature extractor training set (50%), the Attention module training set (40%), and the Tuning set (10%). Both the feature extractor (Resnet-18 CNN) model and the attention module were optimised against binary class labels (NHG 1 and 3) to ensure that the model learns high and low-grade patterns despite substantial label noise (reflected by high inter-assessor variability in NHG grade label assignments). The proposed approach implicitly assumes that the NHG grading follows a continuum of morphological changes (1-3), and that NHG2 is the intermediate group with the highest assessment uncertainty and inter-rater variabilities. We, therefore, excluded NHG 2 from the model optimisation. These three sub-splits were stratified on the clinical NHG (Figure 1b).

The attention-based Multiple Instance Learning (MIL) model was considered as the CNN modelling architecture inspired by Lu et al. (17). It consisted of two separate trainable modules: The feature-extractor and the attention module. The feature extractor was trained to learn breast cancer domain-specific tile-level representations and the attention module was trained to aggregate these tile-level representations to whole slide-level prediction scores. Importantly, we specifically used different sub-splits of the training set for Feature extractor optimisation and Attention module optimisation, respectively,

#### Feature-extractor module

The feature-extractor was a binary weakly-supervised learning model (8,13). We applied the Resnet-18 (18) CNN architecture initialised with weights pre-trained from Imagenet (19). In order to reduce overfitting, we included a dropout layer with a probability of 0.2 after the global average pooling layer. Furthermore, a fully connected layer of 1024 hidden units followed by ReLU activation was added before the final output layer to increase the depth of the architecture. This model was trained on binary labels of NHG 1 vs NHG 3 with cross-entropy loss. We used SGD optimiser (20) with a learning rate of 1e-5 and a momentum value of 0.9. At each training partial epoch end, we used the tuning set to validate the training performance and save the best model according to the lowest validation loss from the tuning set. We applied an early stopping to terminate the training when the validation loss showed no improvement after 50 consecutive partial epochs.

#### Attention module

We used the feature extractor to extract a 512-dimensional feature vector from the average pooling layer for each image tile in the attention-module training set and the tuning set (Figure 1b). These learned features were further used to train the attention module. The attention module consisted of an attention backbone and a classification layer with two output neurons, one for each class (17). The attention backbone assigns and optimises the weights for each tile-level feature vector from each WSI and these derived weights sum up to one in order to be invariant to the number of tiles in each slide. The tile-to-slide feature aggregation was facilitated by the weighted average feature vectors from all image tiles in each slide (21). The attention module was trained as a binary classification task to predict NHG 1 vs NHG 3 tumours using the cross-entropy loss. We used SGD optimiser with a learning rate 1e-5 and a momentum of 0.9. At each training epoch, we used the batch size of one single slide including all image tiles in it, based on our previous work (22).

#### Assignment of predicted histological grade (i.e. predGrade)

We obtained the slide-level predicted scores (i.e. P[class=NHG3|WSI_i_]) for the entire training set (N=1695 WSIs) from the five-fold CV). We further optimised the two thresholds *θ*_1_, *θ*_2_ on P(class=NHG3|WSI_i_) to generate a three-level predicted grade (i.e. predGrade 1, 2 and 3). The thresholds *θ*_1_, *θ*_2_ were established through an exhaustive search by maximizing the agreement between the clinical NHG and the predicted grades (i.e. predGrade 1, 2 and 3) using Cohen’s Kappa Score (*κ*).

### Assessment of model performance

Performance of predGrade was evaluated in both five-fold CV and in the independent external test set (SCAN-B cohort, n=1262). In performance evaluation in the SCAN-B cohort, the five CV models were treated as base models in an ensemble model, where the five predicted scores of P(class=NHG3|WSI_i_) were aggregated using the median across all base-model predictions. Next, we applied the thresholds *θ*_1_*,θ*_2_ (see above) to map predictions to predGrade 1, 2 and 3.

First, we assessed the agreement between the predGrade and the clinical NHG in the independent external test using confusion matrices. Since the clinical NHG has high inter-rater variability, we utilise patient outcome (recurrence-free survival (RFS)), as our primary evaluation metric. We compared the prognostic performance of predGrade and the clinical NHG grade. The RFS defined recurrence (i.e local or distant metastasis, detection of contralateral tumours) or death as the event outcome. Patients were followed from the initial diagnosis to the date of death/recurrence, emigration, or the last registration date, whichever occurred first. Kaplan-Meier (KM) curves for the predGrade and the clinical NHG on RFS using time since the initial diagnosis as the underlying time scale was used for visualisation purposes. Differences in survival probability among clinical NHG and predGrade subgroups were tested using the log-rank test. We assessed the associations between predGrade and RFS as well as clinical NHG and RFS separately by estimating hazard ratios (HRs) with 95% confidence intervals (CIs) using the Cox Proportional Hazard (PH) models. We used the time since the initial diagnosis as the underlying time scale. First, we fitted univariate Cox models for the predGrade and clinical NHG, respectively. Next, we fitted multivariable Cox models additionally adjusting for the well-established clinicopathological factors including tumour size, ER status, HER2 status, lymph node status, and age at the diagnosis. Tumour size was dichotomized as *≥20mm or* <20mm. ER status was positive if the immunohistochemical (IHC) staining indicated the presence of more than 10% ER positively stained cells. HER2 status was determined using IHC staining and FISH or SISH assay. Lymph node status denoted the presence of lymph node metastasis. Cases with missingness in the outcome or in any covariate were excluded from analyses. Two-sided alpha of 0.05 was used for all the statistical tests. Statistical testing for differences in hazard ratio estimates between the grade models was performed using the hr.comp2 function in the survcomp package in R (23).

## Results

### Model performance

#### Classification performance in five-fold CV

Classification performance (Figure 3) of predGrade in comparison with clinical NHG was first assessed using CV (see Methods section), indicating moderate agreement between predGrade and clinical NHG (Cohen’s *κ* = 0.33). 4.8% of clinical NHG 1 was classified as predGrade 3, and 5.3% of clinical NHG 3 was classified as predGrade 1.

**Figure 3:**
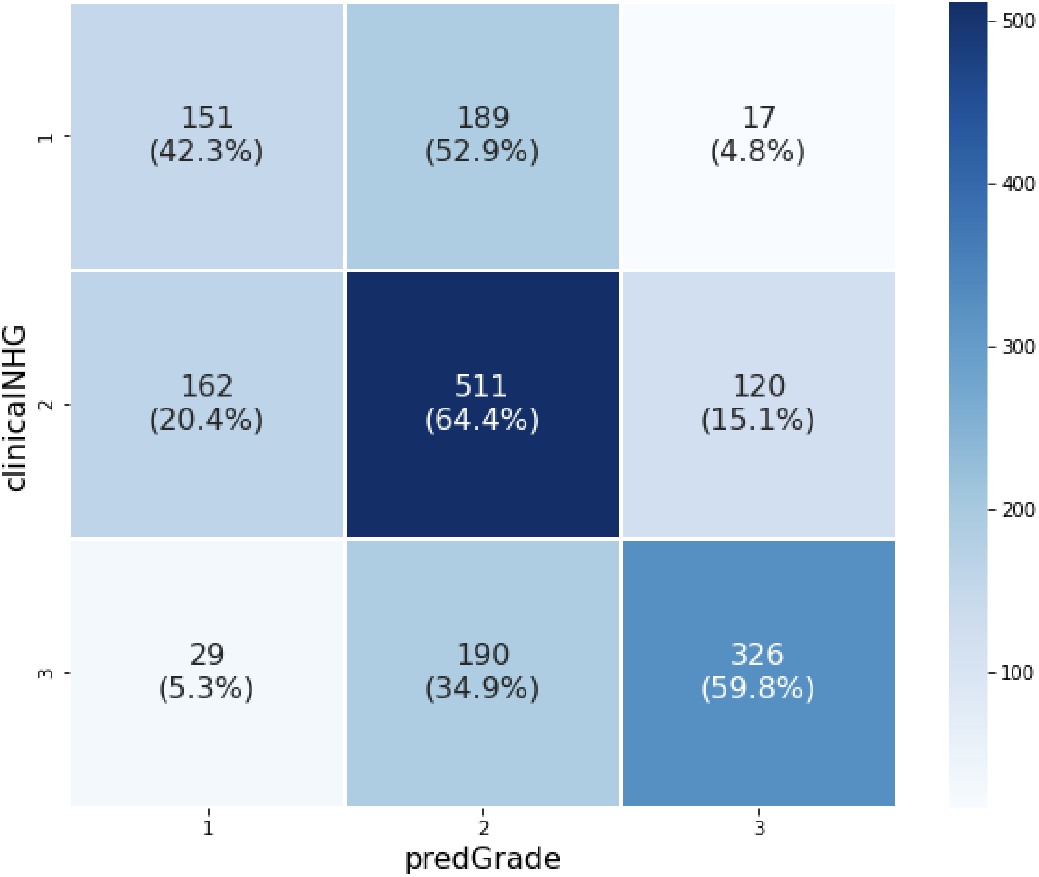
Confusion matrix shows the agreement between the predGrade and clinicalNHG in the five-fold cross-validation.

#### Prognostic performance on RFS in the five-fold CV

Subsequently we evaluated the prognostic performance through time-to-event analysis. Figure 3 showed the KM curves comparing the risk stratification on RFS by clinical and predGrade. We observed similar stratification effects by the predGrade and clinicalNHG. Patients with clinicalNHG 1 or predGrade 1 showed the best survival while patients assigned the clinical NHG 3 or predGrade 3 showed the worst survival (Figure 4).

**Figure 4:**
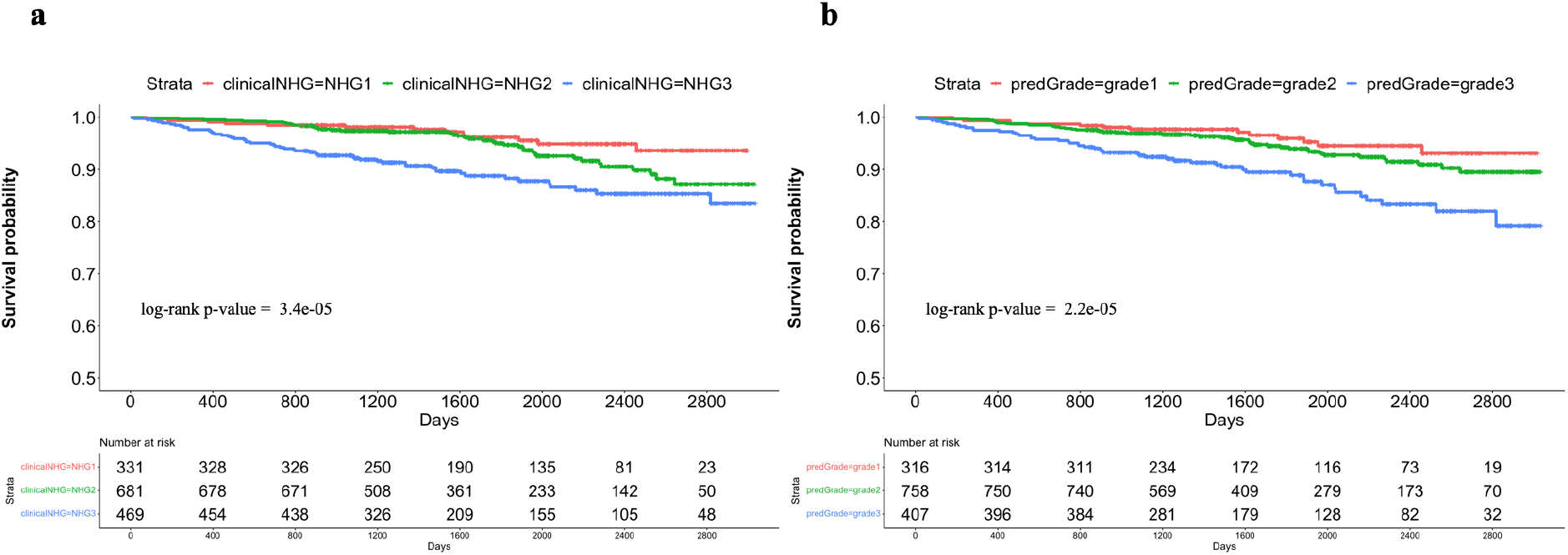
Kaplan-Meier (KM) curves on recurrence-free survival from cross-validation. **a**. KM curve stratified by clinical NHG and **b**. KM curve stratified by predGrade.

In the univariate Cox models, we observed similar effect sizes between the predGrade or clinical NHG and RFS (Figures 5a and 5b). The predGrade 3 (HR= 3.12, 95%CI = 1.69-5.76, p-value < 0.001) and clinicalNHG 3 (HR=3.19, 95% CI =1.74-5.85, p-value < 0.001) showed approximately two times higher risk of an event as compared to predGrade 1 and clinicalNHG 1, respectively. Neither HR estimates for predGrade 2 nor clinicalNHG 2 were found to be significant (p-value < 0.05) (Figure 4a and 4b).

**Figure 5:**
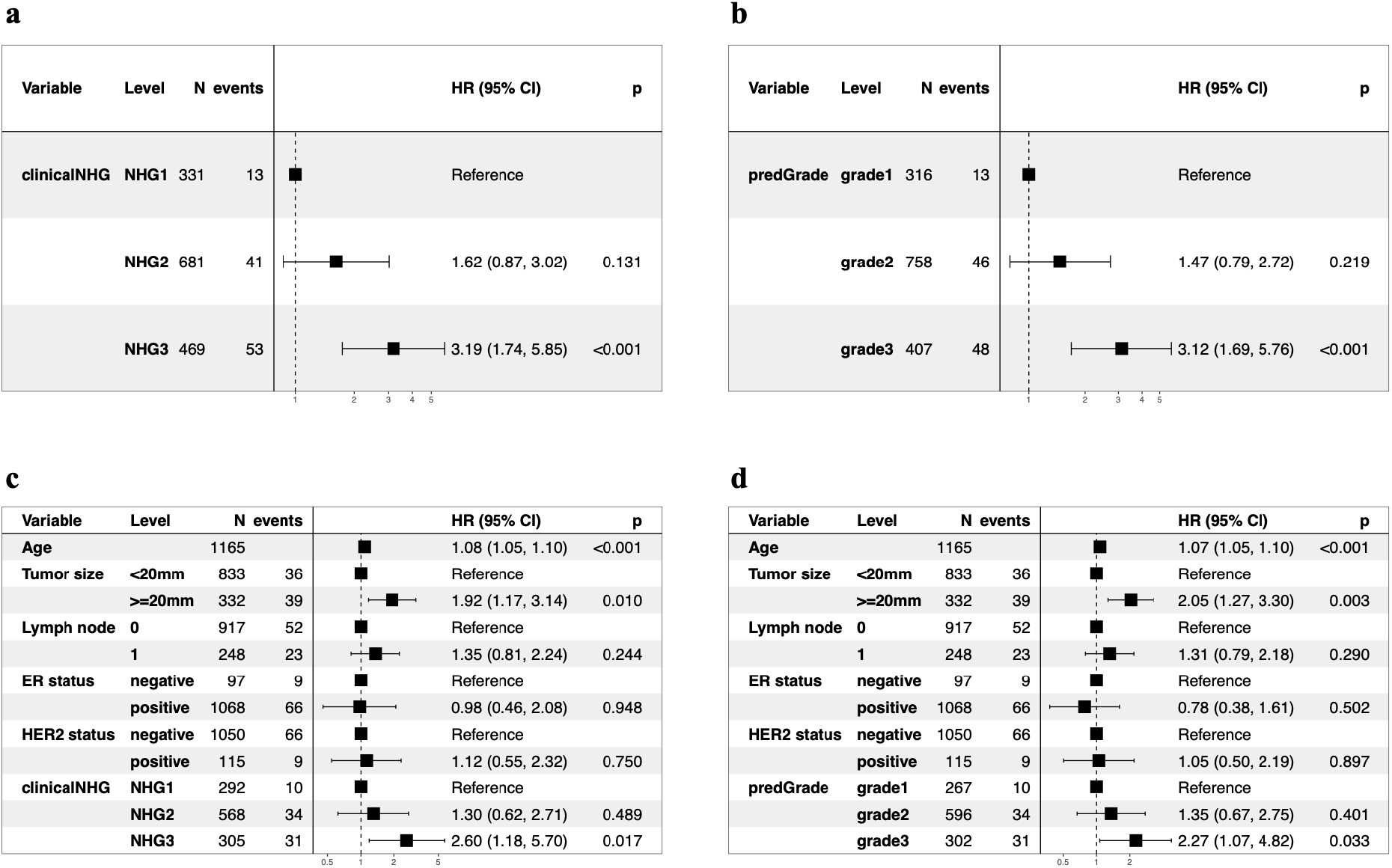
Evaluation of the prognostic performance on recurrence-free survival (RFS) in five-fold CV. **a**. Univariate Cox PH regression analysis between the clinical NHG and RFS; **b**. Univariate Cox PH model between the predGrade and RFS **c**. Multivariable Cox PH model between the clinical NHG and RFS adjusting for age, tumour size, lymph node, ER and HER2 status; **d**. Multivariable Cox PH model between the predGrade and RFS adjusting for age, tumour size, lymph node, ER and HER2 status.

In the multivariable Cox PH models, adjusting for tumour size, lymph node, ER, and HER2 status, the predGrade3 remained associated with a higher risk of death/recurrence (HR=2.27, 95%CI= 1.07-4.82, p-value = 0.033) (Figure 4d). A similar association was also noted for the clinicalNHG 3 (HR= 2.60, 95% CI = 1.18-5.70, p-value = 0.017) (Figure 4c). Neither predGrade2 nor clinicalNHG2 was found to be significantly (p-value < 0.05) associated with RFS (Figures 4c and 4d). In addition, we observed a higher risk of death/recurrence linked to older age and tumour size equal to or larger than 20mm, while the number of lymph nodes, ER status, and HER2 status was not related to RFS (Figure 4c and 4d).

### Model performance in the independent external test set

#### Classification performance in the independent external test set

The classification performance, as assessed by the confusion matrix (Figure 6) and estimation of Cohen’s *κ* = 0.33, were found to be consistent with CV results.

**Figure 6:**
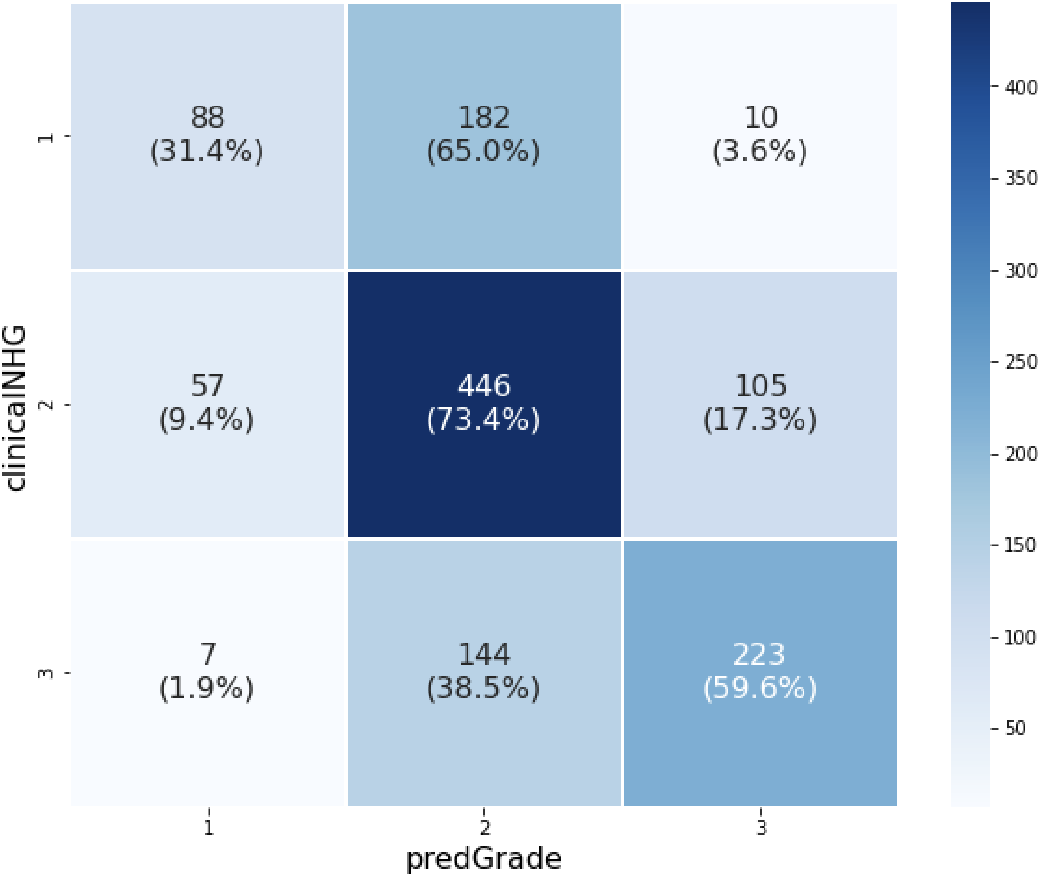
Confusion matrix shows the agreement between the predGrade and clinical NHG in the independent test set.

#### Prognostic performance in the independent external test set

In the independent external test set, KM curves showed similar risk stratification on RFS by the predGrade (log-rank p-value = 0.0002) as compared to the clinical NHG (log-rank p-value = 0.00049) (Figure 7).

**Figure 7:**
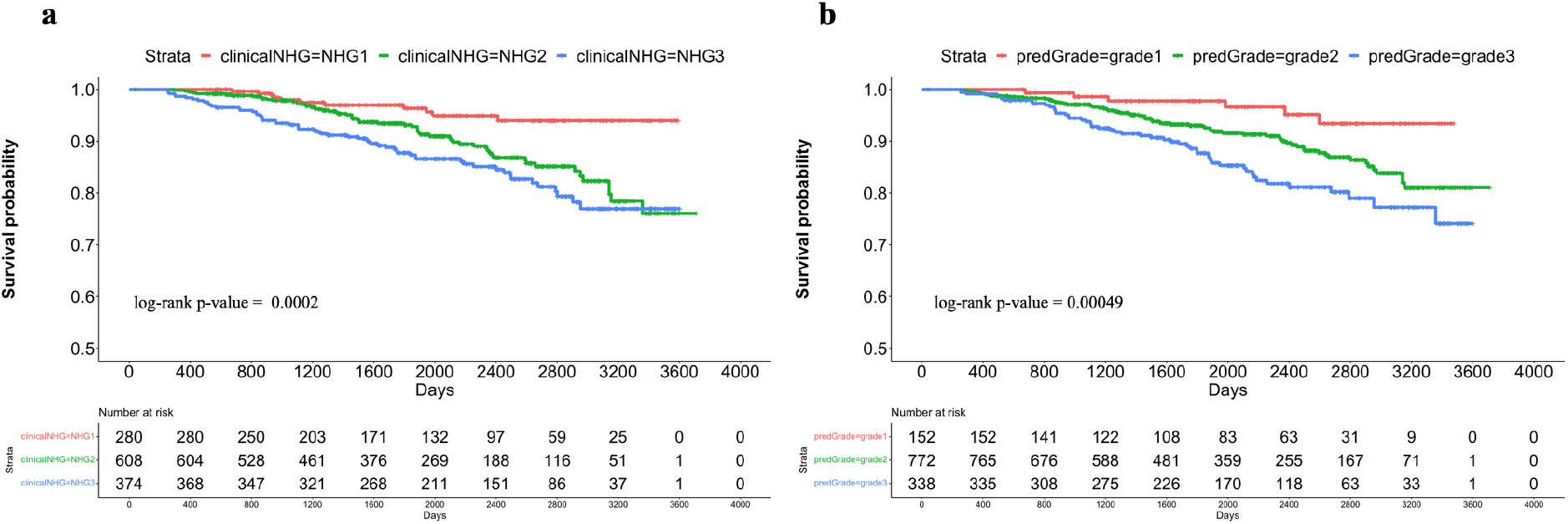
Kaplan-Meier (KM) curves on recurrence-free survival stratified by clinical NHG and predGrade in the independent external test set. **a**. KM curve stratified by the clinical NHG and **b**. KM curve stratified by the predGrade

In the univariate Cox PH model, we observed similar effect sizes in the associations between clinical NHG,predGrade and RFS (Figures 8a and 8b). Patients with clinical NHG 3 (HR=3.58, 95% CI: 1.88-6.81, p-value < 0.001) or predGrade3 (HR=4.07, 95% CI: 1.75-9.47, p-value < 0.001) had three-to-four-fold increased risks of death/recurrence (Figure 5a and 5b) as compared to those with clinical NHG 1 or predGrade 1. On the other hand, the clinical NHG 2 (HR = 2.59, 95% CI: 1.36-4.92, p-value = 0.004) and predGrade 2 (HR= 2.52, 95% CI: 1.10-5.81, p-value = 0.030) were linked to around 2.5-fold increased risk of death/recurrence (Figure 5a and 5b).

**Figure 8:**
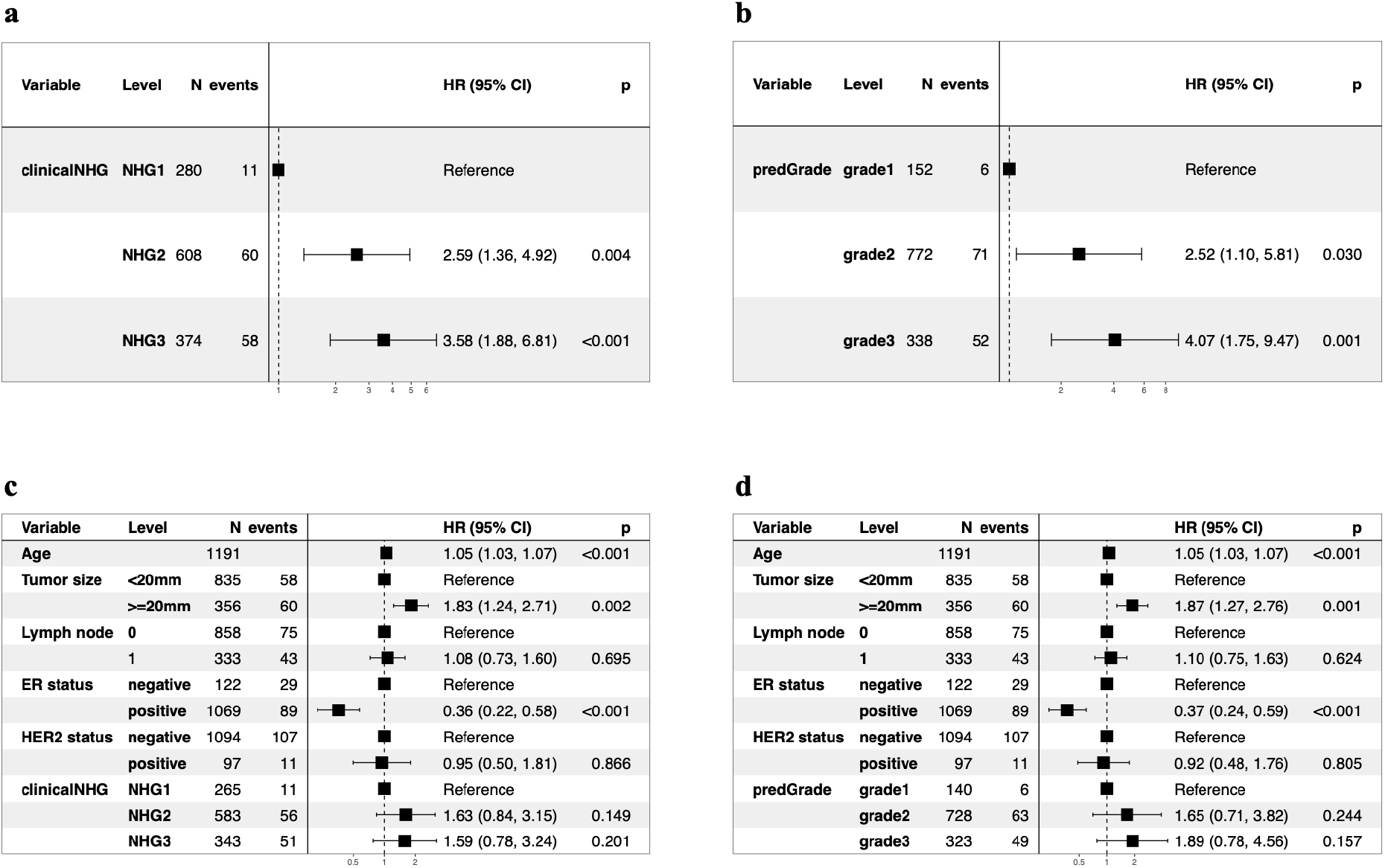
Evaluation of the prognostic performance on recurrence-free survival (RFS) in the independent validation set. **a**. Univariate Cox model between the clinical NHG and RFS; **b**. Univariate Cox model between the predGrade and RFS; **c**. Multivariable Cox model between the clinical NHG and RFS adjusting for age, tumour size, lymph node, ER and HER2 status; and **d**. Multivariable Cox model between the predGrade and RFS adjusting for age, tumour size, lymph node, ER and HER2 status.

In multivariable Cox PH analysis, adjusting for tumour size, lymph node, ER, and HER2 status, the associations between the predGrade, as well as the clinical NHG, with RFS were no longer found to be statistically significant (Figure 8c and 8d), while the effect size estimate was in the same direction as for the univariate analysis. In addition, we noted that older age at diagnosis and larger tumour size was linked to a higher risk of death/recurrence, while ER positive was related to a lower risk of death/recurrence (Figures 8c and 8d). The number of lymph nodes and HER2 status was not related to RFS (Figures 8c and 8d).

Next, we tested for the difference in hazard ratio estimates for clinical NHG 2 vs 1 and predGrade 2 vs 1, indicating no statistically significant difference (p-value > 0.05). We, also tested for the difference in hazard ratio estimates for clinical NHG 3 vs 1 and predGrade 3 vs 1, revealing no significant difference (p-value > 0.05). The hazard ratios in this analysis were calculated from the multivariate Cox PH model after adjusting for the established covariates.

### Subgroup analysis restricting to ER (+ve) or ER(+ve)/HER2(-ve) groups in the independent test set

We plotted KM curves on RFS stratified by clinical NHG and predGrade among ER (+ve) patients (Figure 9a and 9b) and ER (+ve)/HER2(-ve) patients (Figure 9c and 9d).

**Figure 9:**
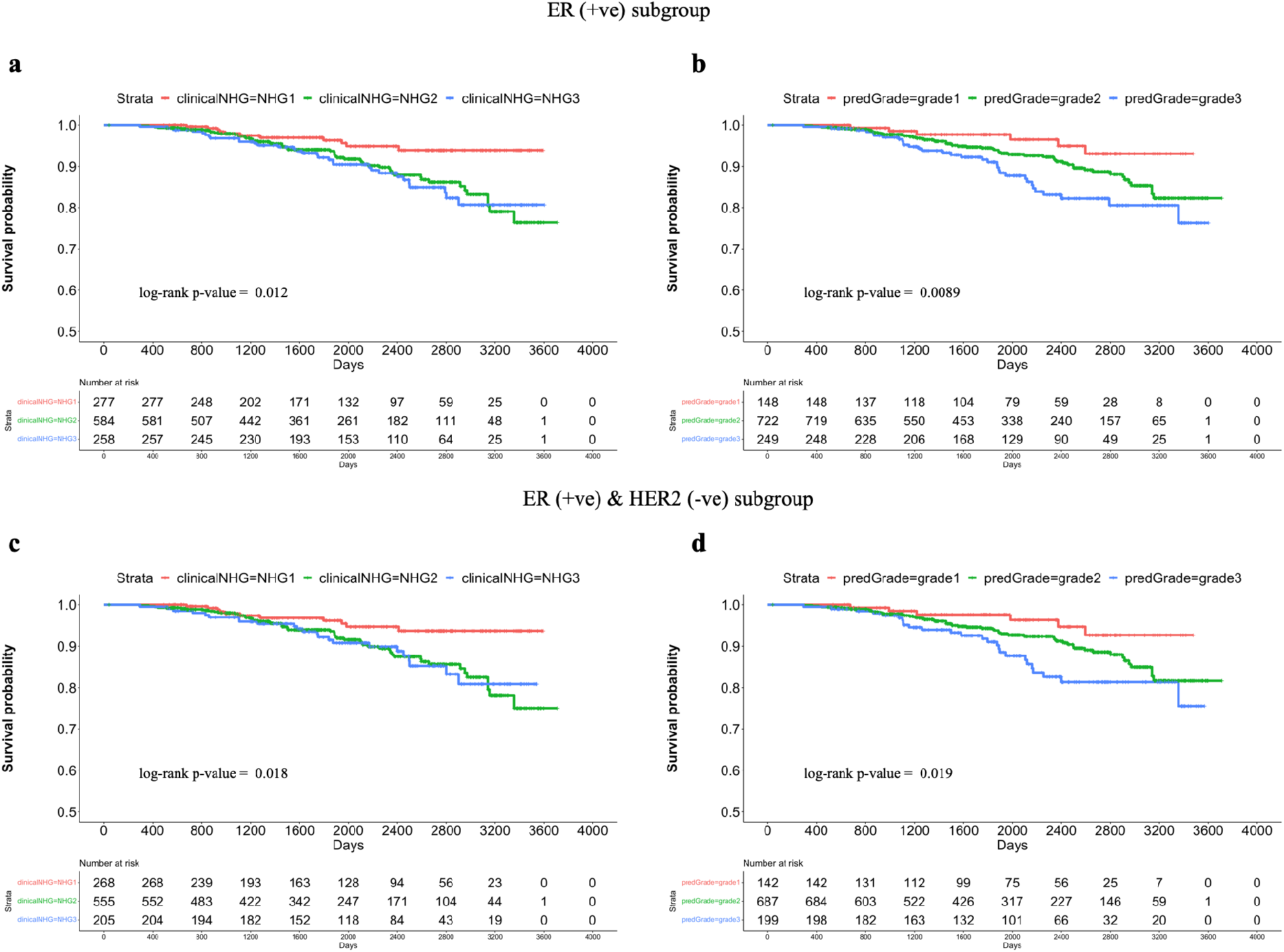
Subgroup analysis: Kaplan-Meier (KM) curves on recurrence-free survival (RFS) in the independent validation set within ER(+ve) or ER(+ve)/HER2(-ve) groups. **a**. KM stratified by clinical NHG in ER(+ve) patients; **b**. KM stratified by predGrade in ER(+ve) patients; **c**. KM stratified by clinical NHG in ER(+ve)/HER2(-ve) patients. **d**. KM stratified by predGrade in ER(+ve)/HER2(-ve) patients

In the univariate Cox model restricted to ER (+ve) patients, we observed increased risks of death/recurrence associated with clinical NHG 3 (HR =2.63, 95% CI: 1.32-5.24, p-value = 0.006) or predGrade 3 (HR = 3.27, (95% CI:1.37-7.82, p-value = 0.008) (Figure 10a and 10b). The clinicalNHG 2 had a 2.42-fold increased risk of death/recurrence, while the predGrade2 was not related to RFS albeit a similar point estimate HR of 2.07 (95% CI: 0.86-4.64, p-value = 0.091) (Figure 10a and 10b). In the multivariable analysis, the association between clinical NHG and RFS as well as the association between predGrade and RFS diminished and were no longer statistically significant (Figure 10 c and 10d).

**Figure 10:**
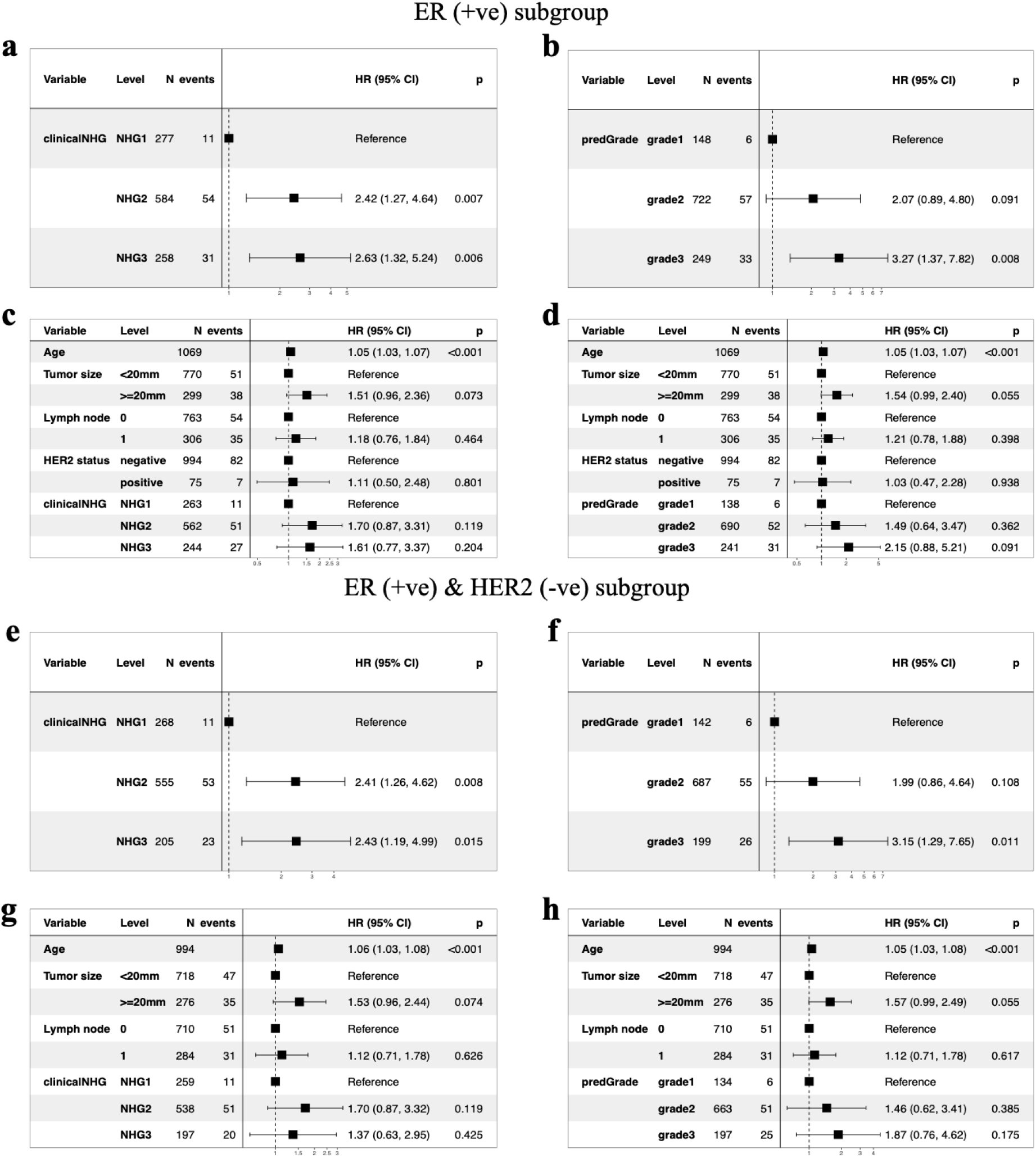
Evaluation of the prognostic performance (RFS) of predGradeon in the independent external test cohort within ER(+ve) or ER(+ve)/HER2(-ve) groups. Univariate Cox PH model for **a**. clinical NHG and **b**. predGrade on RFS among ER(+ve) patients, **c**. Multivariable Cox PH models between c) clinical NHG, **d**. predGrade and FRS among ER(+ve) patients adjusting for age, tumour size, and lymph node. Univariate Cox PH model between **e**. clinical NHG, **f**. predGrade and RFS among ER(+ve)/HER2(-ve) patients. Multivariable Cox model between **g**. clinical NHG, **h**. predGrade and RFS among ER(+ve)/HER2(-ve) patients adjusting for age, tumour size, and lymph node.

Among the ER(+ve)/HER2(-ve) patients, we observed HR = 2.43 (95% CI: 1.19-4.99, p-value = 0.015) for clinical NHG 3 and HR = 3.15 (95% CI: 1.29-7.65, p-value = 0.011) for predGrade 3 (Figure 10e and 10f). The clinical NHG 2 was associated with the RFS (HR=2.41, 95%: CI 1.26-4.62, p-value = 0.008), while the predGrade2 was not related to the RFS (Figures 10e and 10f). In the multivariable Cox PH models among ER(+ve)/HER2(-ve) patients, neither clinical NHG nor predGrade were related to RFS (Figure 10g and 10h), while older age was linked to poor RFS in the analysis for predGrade (Figure 10h).

Again, we tested for the difference in HR estimates between NHG and predGrade in the subgroup analyses, both for grade 1 vs 2 and grade 1 vs 3, and found that neither was significantly different (p-value > 0.05), indicating that the prognostic performance was similar between NHG and the predGrade model.

## Discussion

In this study, we developed a MIL-based CNN model to reproduce clinical NHG breast cancer patients. The proposed model was first evaluated using CV, followed by validation in a fully independent external test set. Histological grading of breast tumours is routinely assessed in the clinical setting and remains an important prognostic factor contributing to clinical decision-making, especially for ER (+ve)/Her2(-ve) patients. However, it is well-known that NHG suffers from substantial inter-assessor and inter-lab variability, which motivates the development of decision-support solutions that can improve quality and consistency in the assessment.

Our proposed model, predGrade, exhibited a moderate label agreement with the clinical NHG (*κ* = 0.33). This imperfect agreement is likely driven by the ground truth labelling noise, both during training and validation, mostly from the intermediate NHG 2, given the high inter-rater variability observed in NHG 2 (3). However, interestingly we observed similar prognostic performances (RFS) for predGrade compared with the clinicalNHG. We also noted similar prognostic performance between predGrade and clinical NHG when restricted to the clinically relevant subgroups of ER(+ve) or ER(+ve) and HER2(-ve) patients. Our results suggest that the deep learning-based predGrade provides similar prognostic performance (RFS) of the clinical NHG, which is a key consideration since the conventional NHG grade is primarily used in clinical settings as a prognostic factor (24). This indicates that deep learning-based solutions can provide decision support based on the same principles of histological grading while offering the benefits of being objective and consistent. The model has the potential to reduce inter-assessor variability between pathologists and systematic variability between pathology labs, which has recently been shown to result in (6) unequal diagnostic quality for patients.

Previous studies have focused on classifying NHG1 and 2 (low-intermediate) combined versus NHG3 (high) (10,11). Wetstein et al. reported a 37% increased risk of recurrence associated with a high grade compared to a low-intermediate grade. Wang et al. on the other hand demonstrated that a deep learning model optimised to discriminate NHG 3 vs 1 can (13) further stratify the intermediate NHG 2 into NHG2-low and NHG2-high, enabling improved prognostic stratification of the NHG 2 group of patients. However, the possibility of utilising a deep learning-based model to reproduce the three-level grade that resembles the conventional clinical NHG has not previously been reported.

In our modelling strategy, we make some key assumptions. Due to the inter-observer and inter-lab variability present in the clinical NHG with relatively higher variability in NHG 2, thus we decided to optimise our model for the classification of NHG 3 and 1 where there is less label noise. As stated earlier, this modelling strategy is based on the assumption that the tumour grading exists in a continuum instead of the discrete labels at the morphological level with a spectrum ranging from low to high grade, apart from assuming more reliable ground truth for low and high NHG. Such variabilities in the ground truth labels are one of the important challenges in developing deep learning-based clinical decision support tools. Especially the development of weakly-supervised learning-based models where the label is only available at the WSI level. An alternative approach to the modelling problem would be to attempt to reduce label noise, which could be achieved by e.g. utilising consensus labels assigned by a set of assessors as performed in (12). However, such attempts remain challenging due to the number of resources required and the shortage of pathologists available in most parts of the world.

In this study we developed and validated a deep learning-based model for breast cancer histological grading, providing a similar three-group grade assignment as the well-established Nottingham Histological Grading system. We found that despite the relatively low concordance of grade labels with clinical NHG, the proposed model provides equivalent prognostic stratification of breast cancer patients. The proposed model has the potential to provide objective and consistent decision support for histological grading, reducing previously observed inter-assessor and systematic inter-lab variability in breast cancer histological grading, and with the benefit of increased equality for patients and reduced risk for over- and under-treatment.

## Data Availability

The data used in the present study is not publicly available

